# Anandamide is an Early Blood Biomarker of Hermansky-Pudlak Syndrome Pulmonary Fibrosis

**DOI:** 10.1101/2024.05.16.24307300

**Authors:** Resat Cinar, Abhishek Basu, Muhammad Arif, Joshua K. Park, Charles N. Zawatsky, Ben Long G. Zuo, Mei Xing G. Zuo, Kevin J. O’Brien, Molly Behan, Wendy Introne, Malliga R. Iyer, William A. Gahl, May Christine V. Malicdan, Bernadette R. Gochuico

**Author notes:** **Corresponding author:** Resat Cinar, Pharm, PhD, MBA, Section on Fibrotic Disorders, National Institute on Alcohol Abuse and Alcoholism, National Institutes of Health, 5625 Fishers Lane, Rockville, MD 20852.

## Abstract

Hermansky-Pudlak syndrome (HPS) is a group of rare genetic disorders, with several subtypes leading to fatal adult-onset pulmonary fibrosis (PF) and no effective treatment. Circulating biomarkers detecting early PF have not been identified. We investigated whether endocannabinoids could serve as blood biomarkers of PF in HPS. We measured endocannabinoids in the serum of HPS, IPF, and healthy human subjects and in a mouse model of HPSPF. Pulmonary function tests (PFT) were correlated with endocannabinoid measurements. In a pale ear mouse model of bleomycin-induced HPSPF, serum endocannabinoid levels were measured with and without treatment with zevaquenabant (MRI-1867), a peripheral CB_1_R and iNOS antagonist. In three separate cohorts, circulating anandamide levels were increased in HPS-1 patients with or without PF, compared to healthy volunteers. This increase was not observed in IPF patients or in HPS-3 patients, who do not have PF. Circulating anandamide (AEA) levels were negatively correlated with PFT. Furthermore, a longitudinal study over the course of 5-14 years with HPS-1 patients indicated that circulating AEA levels begin to increase with the fibrotic lung process even at the subclinical stages of HPSPF. In pale ear mice with bleomycin-induced HpsPF, serum AEA levels were significantly increased in the earliest stages of PF and remained elevated at a later fibrotic stage. Zevaquenabant treatment reduced the increased AEA levels and attenuated progression in bleomycin-induced HpsPF. Circulating AEA may be a prognostic blood biomarker for PF in HPS-1 patients. Further studies are indicated to evaluate endocannabinoids as potential surrogate biomarkers in progressive fibrotic lung diseases.

## Introduction

Hermansky-Pudlak syndrome (HPS) is a rare genetic disorder with 11 reported genetic types, caused by bi-allelic mutations in any of 11 different genes (1, 2). HPS-1 genotype is the most common and severe form. To date, 1,464 individuals with HPS are registered with the HPS Network (www.hpsnetwork.org; personal communication of Donna Appell, R.N.). A major complication in HPS patients is HPS pulmonary fibrosis (HPSPF), which remains a leading cause of mortality with no approved treatment to date (3, 4). HPSPF particularly poses a risk to middle-aged adults with HPS-1 or HPS-4 and children with HPS-2 or HPS-10. Clinically HPSPF shares similarities with idiopathic pulmonary fibrosis (IPF) (5), though unlike IPF, which typically affects older patients and occurs sporadically, HPSPF is highly predictable among adults with HPS-1 based on the natural history of disease (6). Importantly, fibrotic lung disease is progressive in HPS-1, and the identification of biomarkers capturing this progression could allow the initiation of treatment at the earliest stages of the disease.

Endocannabinoids are bioactive lipids that act on cannabinoid receptor 1 (CB_1_R) and cannabinoid receptor 2 (CB_2_R). Cannabinoid receptors also recognize and mediate the effects of the active ingredient of cannabis (7). Anandamide (arachidonoyl ethanolamide, AEA) and 2-arachidonoylglycerol (2-AG) are two well-characterized endocannabinoids that serve as endogenous agonists of the cannabinoid receptors. Endocannabinoids, particularly through CB_1_R promote fibrosis in multiple peripheral organs including liver (8–12), kidney (13–15), heart (16, 17), and skin (18–20). Consequently, peripheral CB_1_R antagonism has emerged as a potential therapeutic strategy for fibrotic disorders, as it has shown efficacy in attenuating organ fibrosis in multiple experimental models (21). Importantly, overactivity of the endocannabinoid/CB_1_R system contributes to pulmonary fibrosis with different etiologies such as radiation-induced PF (22), IPF (23) and HPSPF (24). Notably, elevated AEA levels in the bronchoalveolar lavage fluid (BALF) of IPF (25) and HPSPF (24) patients were found to inversely correlate with pulmonary function test (PFT) parameters, suggesting that AEA could be a biomarker of PF progression in diseases such as HPSPF and IPF.

Endocannabinoids are synthesized and utilized on-demand by multiple cell types in tissues and blood to modulate the cannabinoid receptors that serve autocrine or paracrine functions (26). Dysregulation of the endocannabinoid system in peripheral organs accompanies inflammatory and profibrotic conditions (21). In this study, we investigated endocannabinoids in blood specimens of HPS patients with and without PF. We examined longitudinal samples from patients exhibiting progressive HPSPF and utilized an animal model of HPSPF to further our understanding. Our findings point to circulating AEA in peripheral blood as a promising early biomarker for HPSPF, reflecting the broader role of the endocannabinoid system in regulating cellular and metabolic homeostasis.

## Materials and Methods

### Study Participants and Human Subject Consent

Patients with IPF, HPS and healthy research volunteers provided written informed consent and enrolled in protocol 95-HG-0193 (clinicaltrials.gov NCT00001456, “Clinical and Basic Investigations into Hermansky-Pudlak syndrome”) and/or 04-HG-0211 (clinicaltrials.gov NCT00084305, “Procurement and Analysis of Specimens from Individuals with Pulmonary Fibrosis”), which were approved by the Institutional Review Board of the National Human Genome Research Institute from the National Institutes of Health. Study eligibility criteria were previously described (27).

### Patients

We conducted peripheral blood biomarker analysis in three separate sets of samples from the natural history cohort referred to as the study [1], validation [2], or progression [3] cohort. Blood was collected when patients visited the NIH clinical center and processed to isolate serum according to standard methods. The study cohort was comprised of 90 serum samples from 12 normal volunteers (NVs), 11 HPS-1 patients without PF, 40 HPS-1 patients with HPSPF, 10 IPF patients, and 10 HPS-3 patients without PF (Table 1, Figure 1). The validation cohort comprised of 40 serum samples from five NVs, 10 HPS-1 patients without PF, 10 HPS-1 patients with HPSPF, 5 IPF patients, and 10 HPS-3 or HPS-5 patients without PF (Table 2, Figure 2). To monitor longitudinal, progressive changes of PF in the same HPS-1 patients, we also employed a progression cohort comprised of eight HPS-1 patients (four female and four male) ranging from 35 to 60 years of age. Each patient had five or six serial blood samples, PFTs and HRCT imaging during a longitudinal follow up ranging from 5 to 14 years.

**Figure 1:**
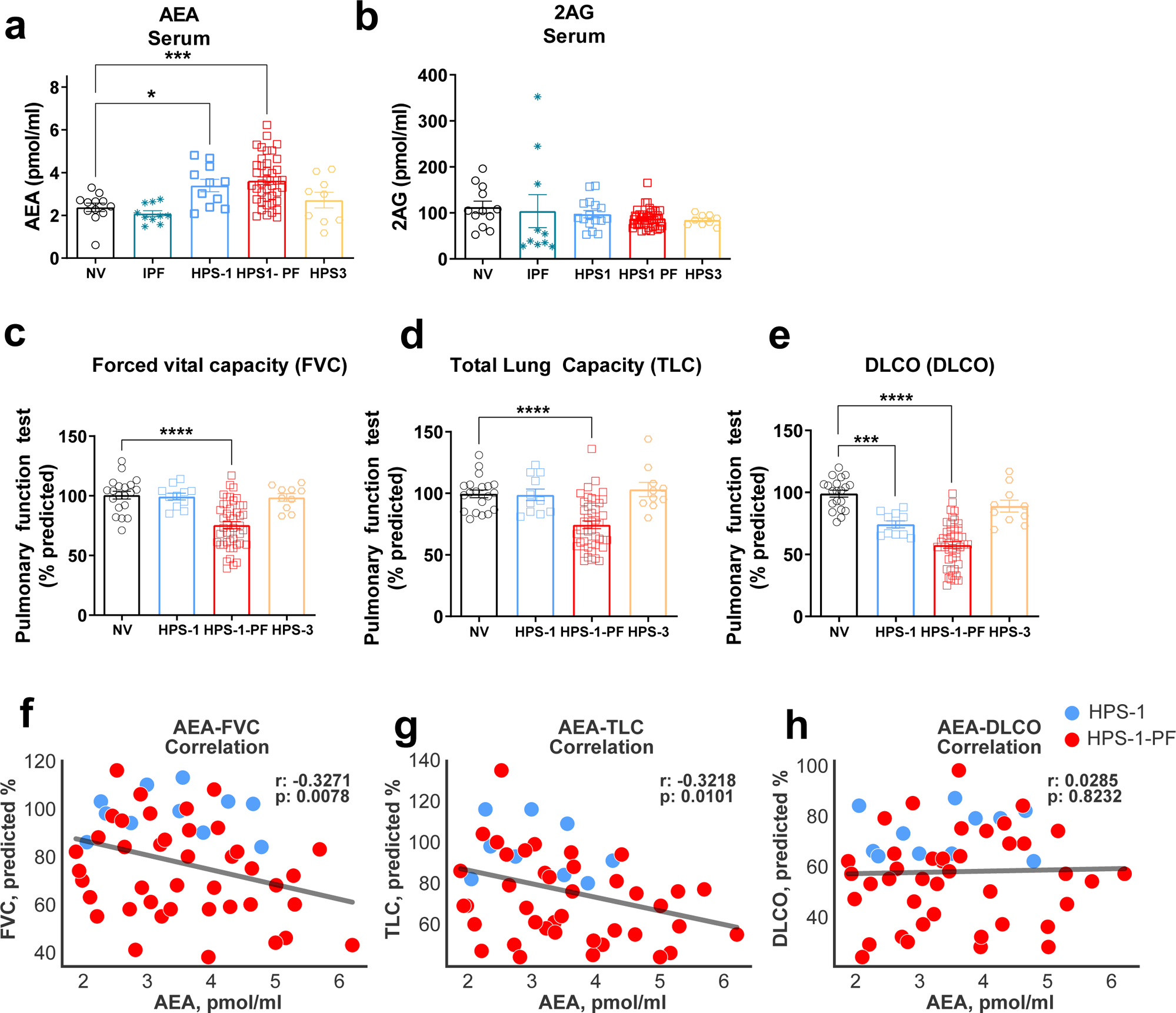
Increased level of Anandamide in serum of HPS1 patients was negatively correlated with pulmonary function test parameters. Levels of endocannabinoid AEA (**A**) and 2AG (**B**) in serum from normal volunteers (NV), HPS-1 without fibrosis, patients with HPSPF, HPS-3 patients and IPF patients. Pulmonary function tests (PFTs) in the same groups. FVC = forced vital capacity (**C**), TLC = total lung capacity (**D**), DLCO = diffusion capacity (**E**). Correlation with PFTs and AEA in serum in HPS-1 and HPSPF (**F, G, H**), HPS1: blue symbol, HPSPF: red symbol. Correlation was calculated by using Pearson correlation coefficients. Data represent mean + S.E.M. from 12 NV, 11 HPS-1, 40 HPSPF, 10 HPS-3, and 10 IPF subjects for serum and PFTs. Data were analyzed by one-way ANOVA followed by Dunnett’s multiple comparisons test for endocannabinoids and PFT. *(*P* ≤ 0.05), **(*P* ≤ 0.01), ***(*P* ≤ 0.001), ****(*P* ≤ 0.0001).

**Figure 2.**
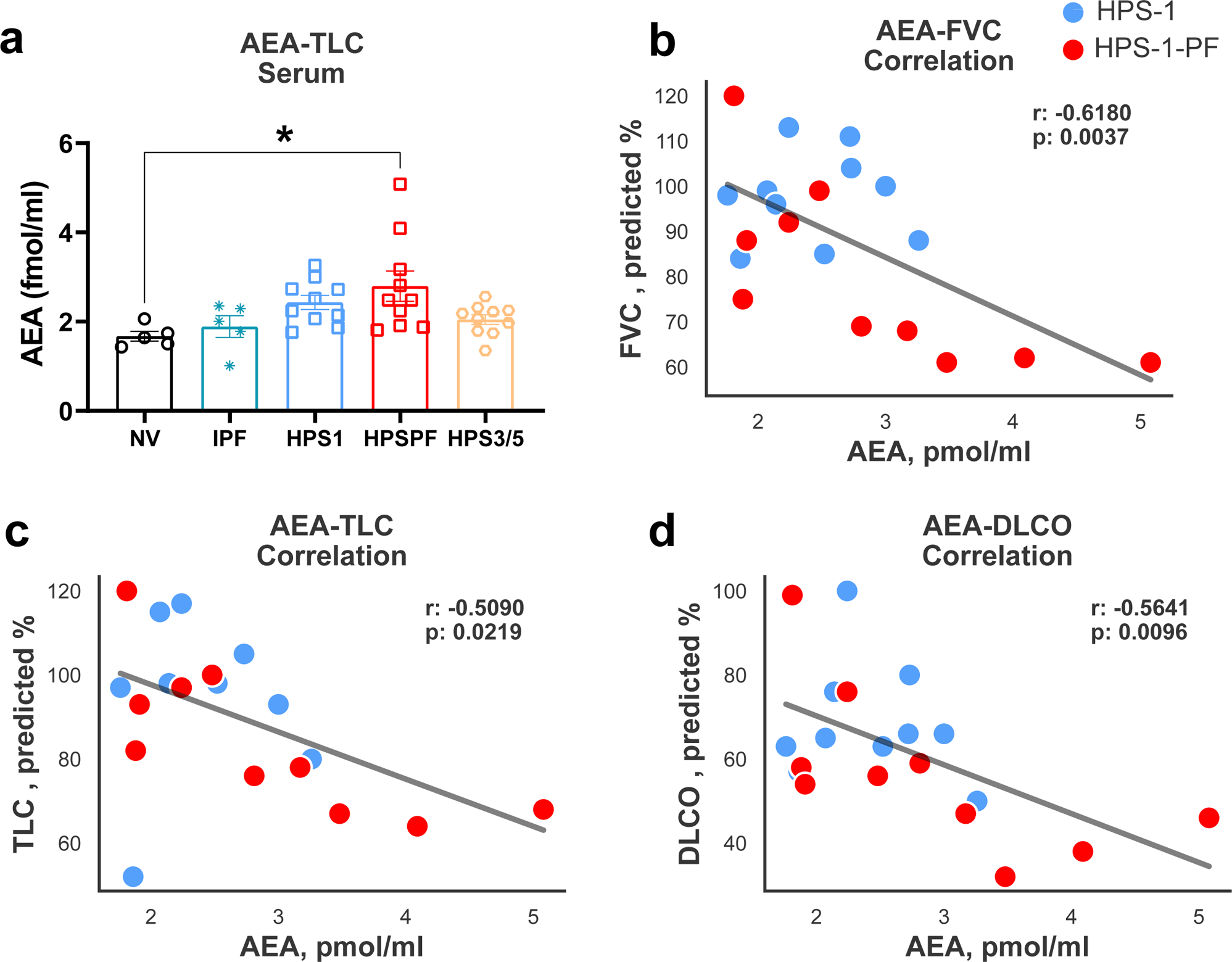
Level of AEA in serum and its correlation with PFTs in the validation cohort subjects. (**A**) Levels of AEA in serum from the patients in a validation cohort consist of normal volunteers (NV), HPS-1 without fibrosis, patients with HPSPF, HPS-3 or HPS-5 patients and IPF patients. Correlation with AEA and PFTs FVC (**B**), TLC (**C**), DLCO (**D**) in serum in HPS-1 and HPSPF subjects, HPS1: blue symbol, HPSPF: red symbol. Correlation was calculated by using Pearson correlation coefficients. Data represent mean + S.E.M. from 5 NV, 10 HPS-1, 10 HPSPF, 10 HPS-3, HPS-5, and 5 IPF subjects for serum and PFTs. Data were analyzed by one-way ANOVA followed by Dunnett’s multiple comparisons test for endocannabinoids. * (*P* < 0.05) indicates significant difference from the NV group.

**Table 1.**
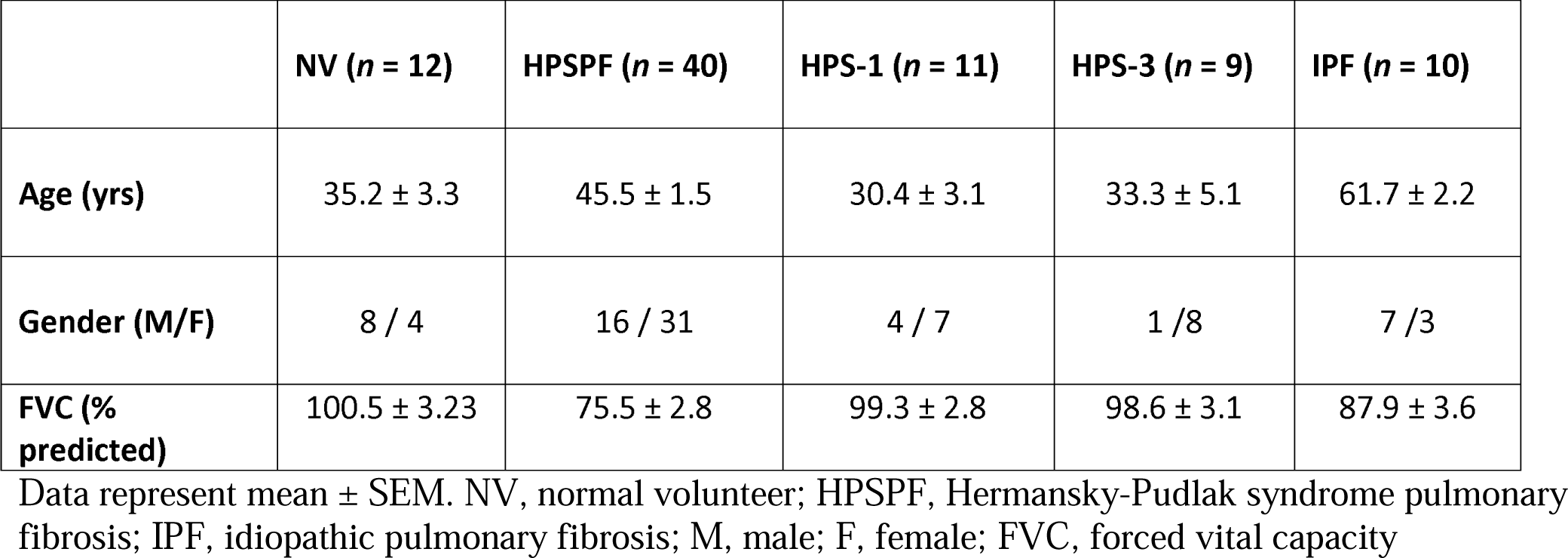
Human Subject Characteristics for Serum Samples in the Study Cohort.

**Table 2.** Human Subject Characteristics for Serum Samples in the Validation Cohort.

|  | NV (*n* = 5) | HPSPF (*n* = 10) | HPS-1 (*n* = 10) | HPS-3 and HPS-5 (*n* = 10) | IPF (*n* = 5) |
| --- | --- | --- | --- | --- | --- |
| Age (yrs) | 41.6 ± 8.28 | 43.7 ± 2.00 | 26.0 ± 2.60 | 34.8 ± 5.83 | 68.8 ± 4.62 |
| Gender (M/F) | 2/3 | 8/2 | 5/5 | 1/9 | 3/2 |
| FVC (% predicted) | 99.2 ± 4.60 | 79.5 ± 6.24 | 97.8 ± 3.17 | 98.5 ± 3.42 | 84.6 ± 12.94 |

Data represent mean ± SEM. NV, normal volunteer; HPSPF, Hermansky-Pudlak syndrome pulmonary fibrosis; IPF, idiopathic pulmonary fibrosis; M, male; F, female; FVC, forced vital capacity

### Pulmonary Function Tests for Human Subjects

PFT parameters were performed as described (28). Briefly, forced vital capacity (FVC), total lung capacity (TLC), and diffusion capacity of carbon monoxide (DLCO) measurements were made in accordance with American Thoracic Society guidelines (SensorMedics, Yorba Linda, CA, USA). Values were expressed as percentages of predicted values.

### High-Resolution Computed Tomography

High-resolution computed tomography (HRCT) scans of the chest without intravenous contrast were performed in the prone position at end-inspiration. Pulmonary fibrosis was diagnosed by characteristic findings on HRCT scans of the chest as previously described (29, 30).

### Chemicals

S-MRI-1867 (zevaquenabant) was synthesized as described (31, 32). Pharmaceutical grade bleomycin was from Hospira (Lake Forest, IL, USA). All the other chemicals were from Sigma–Aldrich (St. Louis, MO, USA).

### Experimental Drug Treatment

Zevaquenabant was administered by oral gavage once daily as indicated and illustrated in Figure 5A. The vehicle was a 1:1:18 ratio of DMSO:Tween 80:Saline. Oral formulations were applied at 1 mg/mL concentrations to achieve dose of 10 mg/kg, which provided the target exposure, the target engagement and antifibrotic efficacy in bleomycin-induced PF models (23, 24).

### Animals

All animal procedures were conducted in accordance with the rules and regulations of the Institutional Animal Care and Use Committee of the National Institutes of Alcohol Abuse and Alcoholism (NIAAA), under the protocols of LPS-GK1. We used male pale ear (*Hps1*^ep/ep^) mouse model since it is the most widely used model of HPS-1, as it mimics structural abnormalities observed in patients with HPS and is susceptible to bleomycin-induced PF (33). Pale ear mice were on a C57Bl/6J genetic background. Mice were housed individually under a 12-hour light/dark cycle and fed a standard diet, *ad libitum* (Teklad NIH-31; Envigo, Huntingdon, UK).

### Subcutaneous Osmotic Delivery of Bleomycin

We generated a bleomycin-induced PF model by delivering bleomycin (60 U/kg/7 days) via subcutaneous osmotic delivery (33, 34) as previously described.

### Endocannabinoid Measurement

Tissue levels of endocannabinoids were measured by stable isotope dilution liquid chromatography/tandem mass spectrometry (LC-MS/MS) as described (31). Briefly, Anandamide (AEA) and 2-Arachidonyl Glycerol (2-AG), were quantified in human serum samples by liquid chromatography/tandem mass spectrometry. 100 µL serum was incubated at −20°C for 10 min with 900 ml ice-cold acetone and 400 µL Tris buffer (50 mM, pH 8.0) to precipitate proteins. After spinning at 2000g at 4°C for 10 min, the supernatant was transferred to a glass tube to evaporate the acetone phase under nitrogen flow. Then, 0.5 mL of ice-cold methanol/Tris buffer (50 mM, pH 8.0) added to the supernatant, 1:1, containing 7 ng of [^2^H_4_] arachidonoyl ethanolamide ([^2^H_4_] AEA), and 50 ng of [^2^H_5_] arachidonoyl glycerol ([^2^H_5_] 2AG), as internal standards. Then, the supernatant was extracted two times with 2 ml of CHCl3:MeOH (2:1, vol/vol). Lower chloroform phase was collected and transfer another glass tube. Then combined chloroform phases were dried under nitrogen flow. The dried samples were reconstituted in 50 µL of ice-cold methanol prior to loading autosampler for mass spectrometry measurements.

LC-MS/MS analyses were conducted on an Agilent 6470 triple quadrupole mass spectrometer (Agilent Technologies) coupled to an Agilent 1200 LC system. Liquid chromatographic separation was obtained using 2 µl injections of samples onto a InfinityLab Poroshell 120 EC-C18 column (3.0mm×100 mm, 2.7 Micron) from the Agilent Technologies. The autosampler temperature was set at 4^◦^C and the column was maintained at 34^◦^C during the analysis. Gradient elution mobile phases consisted of 0.1% formic acid in H_2_O (phase A) and 0.1% formic acid in MeOH (phase B). Gradient elution (350 μL/min) was initiated at 10% B, followed by a linear increase to 50% B at 0.5 min, followed by a linear increase to 85% B at 3 min and maintained until 14 min, then increased linearly to 100% B at 18 min and maintained until 20 min. The mass spectrometer was set for electrospray ionization operated in positive ion mode. The source parameters were as follows: capillary voltage, 3,500 V; gas temperature, 300 °C; drying gas, 5 L/min; nitrogen was used as the nebulizing gas. Collision-induced dissociation was performed using nitrogen. Levels of each compound were analyzed by multiple reaction monitoring. The molecular ion and fragment for each compound were measured as follows: m/z 348.3→62.1 for AEA, m/z 379.3→287.2 for 2AG, m/z 384.3→91.1 for [^2^H_5_] 2AG, m/z 352.3→66.1 for [^2^H_4_] AEA, The Analytes were quantified using MassHunter Workstation LC/QQQ Acquisition and MassHunter Workstation Quantitative Analysis software (Agilent Technologies). Levels of analytes in the samples were measured against standard curves.

### Real-time PCR analysis

RNA extraction and real-time PCR was conducted as previously described (24) using predesigned mouse *Tbp* (QT00198443), *Timp1 (*QT00996282), *Fn1 (*QT00135758), and *Ccl2 (*QT00167832) primers were purchased from Qiagen.

### Fibrosis quantification by hydroxyproline measurements

Lung fibrosis was quantified biochemically by measuring hydroxyproline (Hyp) content of lung extract using LC-MS/MS as previously described (34).

### Statistical Analysis

Statistical analysis was performed by unpaired two-tailed Student’s *t* test or by one-way ANOVA, as appropriate and indicated in figure legends. *P* < 0.05 was considered significant.

## Results

### AEA Levels are Increased in Serum of HPS-1 Patients With or Without PF, but not in IPF Patients

We analyzed serum endocannabinoid levels in healthy normal volunteers (NVs), HPS-1 patients with or without PF, IPF patients, and HPS-3 patients without PF (Table 1, Figure 1). IPF patients served as positive controls for pulmonary fibrosis, whereas HPS-3 subjects were used as nonfibrotic HPS controls because HPS-3 patients do not develop PF (35). AEA levels were significantly elevated only in serum of HPS-1 patients with and without PF, but not in IPF patients, healthy volunteers, or HPS-3 patients without PF (Fig. 1A). Interestingly, 2AG levels remained unchanged across all groups (Fig. 1B). This pattern of AEA elevation in HPS-1 patients suggests specific alteration in their endocannabinoid profile.

### Increased anandamide levels in serum from HPS-1 patients are negatively correlated with pulmonary function tests

In this study cohort, mean FVC was 76 % predicted in HPS-1 patients with HPSPF, which is consistent with their diagnosis of PF. Analyses of PFTs showed that FVC, TLC and DLCO were significantly reduced in HPSPF patients compared to healthy volunteers (Figure 1C-E). To assess a relationship between AEA levels and PFT decline in HPS-1 patients, we analyzed the correlation between serum levels of AEA and pulmonary function parameters among HPS-1, and HPSPF patients (Figure 1F-H). Analyses of PFTs showed that FVC, TLC and DLCO were significantly reduced in HPSPF patients compared to healthy volunteers. Circulating AEA levels were negatively correlated with PFT parameters such as FVC and TLC in human subjects (Figure 1F-H), suggesting that higher concentrations of circulating AEA may contribute to the fibrogenic process in lungs of HPS-1 patients.

We used another group of patients as a validation cohort, comprised of 40 subjects including NV or patients with HPS-1 without PF, HPSPF, HPS-3 or HPS-5, or IPF (Table 1, Figure 2). In the validation cohort, the mean FVC values were 97.8% predicted in the HPS-1 patients without PF and 79.5% predicted in the HPSPF patients. Consistent with the study cohort, circulating AEA levels were higher in the HPS-1 patients with or without PF in this validation cohort. However, this increase was only statistically significant in the HPSPF patients (Figure 2A). Notably, increased AEA levels were negatively correlated with PFT parameters (Figure 2B-D), which validated the observations from the study cohort (Figure 1).

### Identification of Circulating AEA as an Early Disease Prognostic Biomarker in HPSPF

In both the study and the validation sample sets, the circulating AEA levels exhibited a higher trend in HPS-1 patients without PF compared to the NV. Since AEA levels were negatively correlated with pulmonary function decline among HPS-1 subjects, this suggests that AEA could be an early disease biomarker with the increase in HPS-1 subjects at early subclinical stage of PF. HPS-1 patients develop progressive PF with age. Early, subclinical interstitial lung disease can be diagnosed by HRCT lung scans in adults with HPS-1 prior to respiratory symptoms (35). Therefore, we measured circulating AEA levels in the eight HPSPF subjects with progressive disease who had serial blood collections, PFT measurements and HRCT scan over a 5 to 14 years duration of follow up. In this progression cohort, age and circulating AEA levels were increased with the progressive PF in HPS-1 patients (Figure 3A, 3B). Data from three representative patients in the progression cohort show worsening of bilateral ground glass and/or reticulations (Figure 4A-C). The Patient 1 progressed from minimal HPSPF with a FVC of 90% predicted and DLCO of 83% predicted to severe HPSPF with FVC of 43% predicted and DLCO of 20% predicted over 10 years from mid-40’s to mid-50’s (Figure 4A and 4D). The Patient 2 progressed from mild to moderate HPSPF with a decline in FVC from 84 % predicted to 57% predicted in 6 years from mid-40’s to late 40’s (Figure 4B and 4E). The Patient 3 progressed from minimal to severe HPSPF (Figure 4C) with a decline in FVC from 101% to 50% in 10 years from mid-30’s to mid-40’s (Figure 4F). In parallel to the progression of HPSPF in the three patients, the circulating AEA levels were also increased with age and declining pulmonary function (Figure 4D-F). This suggests that circulating AEA levels begin to increase with the fibrotic lung process even at the subclinical stages of HPSPF, and implies its utility as a prognostic marker.

**Figure 3.**
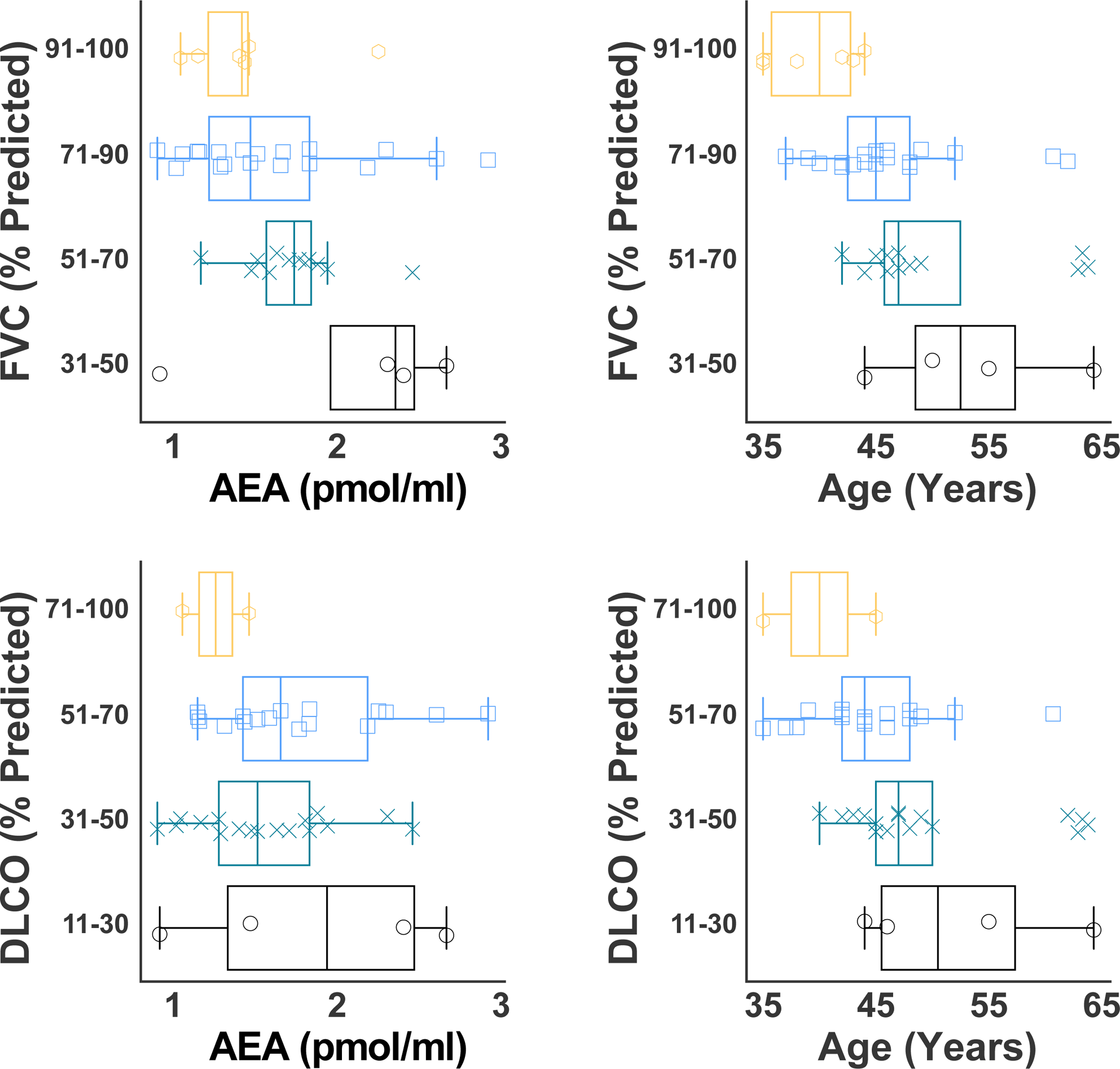
Both circulating AEA and age increase while pulmonary function decline in HPSPF. (**A**) Distribution of age with FVC (**A**) and DLCO (**B**) measurements is shown for a progression cohort. Distribution of circulating AEA level with FVC (**C**) and DLCO (**D**) measurements is displayed. Eight HPS-1 subjects with diagnosis of HPSPF were studied longitudinally to assess progression of pulmonary fibrosis for an interval ranging from 5 to 12 years.

**Figure 4.**
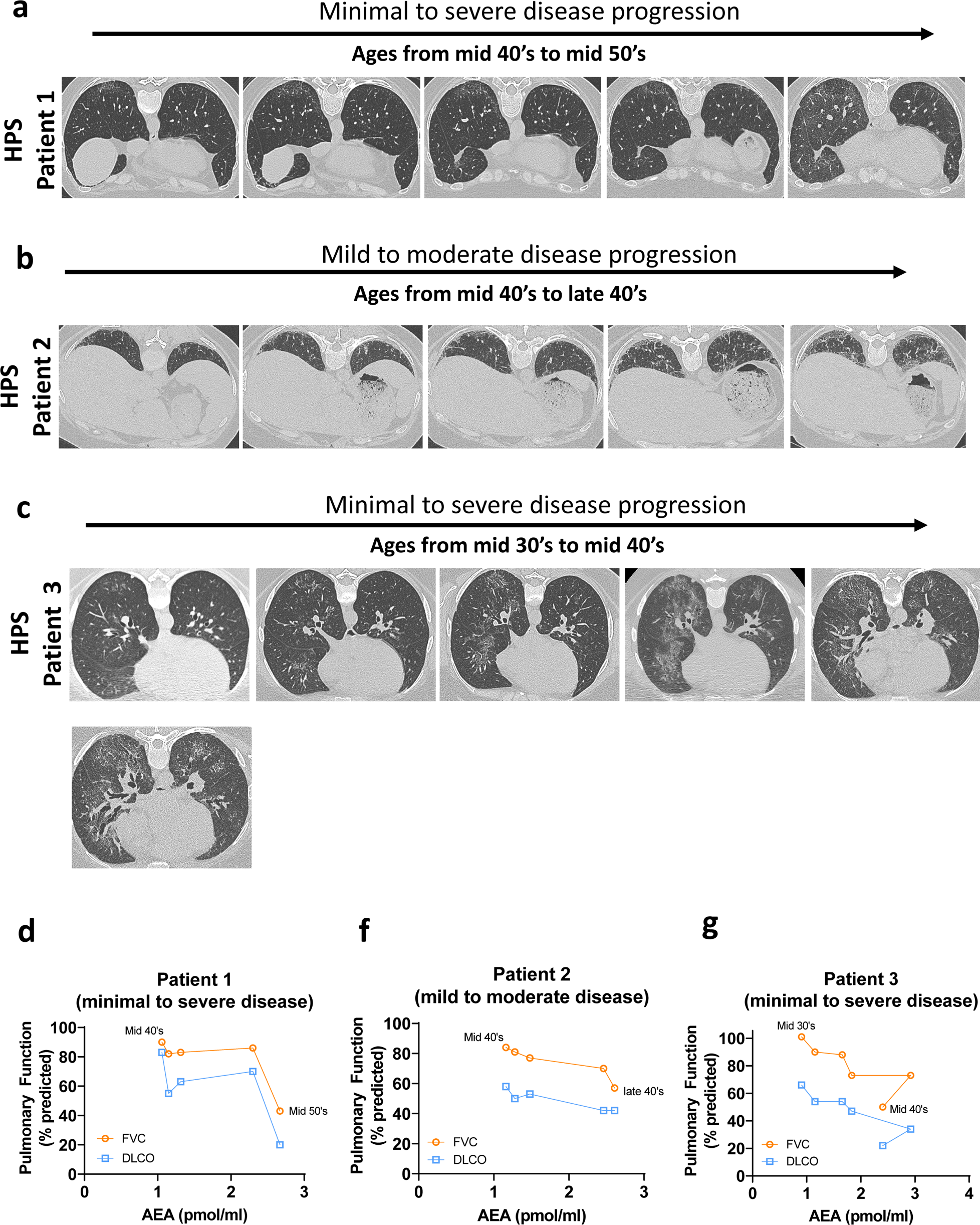
Time dependent changes in circulating AEA levels in HRCT guided progression of HPSPF. Representative HRCT images in HPS patient 1 (**A**), HPS patient 2 (**B**), HPS patient 3 (**C**) who had progressive HPSPF. Age dependent changes in AEA levels in serum in HPS patient 1 (**D**), HPS patient 2 (**E**), HPS patient 3 (**F**).

### Circulating AEA Levels Increased at Subclinical Fibrotic Stage in Bleomycin-Induced HpsPF Model Using Pale Ear Mice

Next, we analyzed the serum of pale ear (*Hps-1*^ep/ep^) mice after bleomycin-induced PF (HpsPF) (Figure 5A), to determine whether circulating AEA increased similarly as in human HPSPF. In this HpsPF model, 8 days after osmotic minipump implantation and bleomycin administration, gene expression for the fibrogenic marker *Col1a* was significantly increased, although no quantifiable fibrosis was observed biochemically or histologically in the previous study (24). This suggests that Day 8 post bleomycin could represent the fibrosis initiation phase in this model since there was no quantifiable fibrosis despite the remarkable increase in the gene expression of fibrosis. Fibrosis was evident 42 days after initial bleomycin treatment (24). Notably, the endocannabinoid/CB_1_R system was overactivated with bleomycin-induced HpsPF in pale ear mice in lungs, as also observed in human HPSPF patients (24), suggesting translational relevance of this model to investigate endocannabinoids. Therefore, this model could also be instrumental to evaluate the status of circulating AEA in subclinical and clinical stages of HPSPF. Same as the previous study, there was no quantifiable fibrosis based on hydroxyproline measurement in lungs at Day 8 post bleomycin (Figure 5B), despite a significant increase in the gene expression of fibrotic markers Timp1 and Fibronectin 1 (Figure 5C) and inflammatory markers Ccl2 (Fig.5d). Similar to results for HPS-1 and HPSPF patients (Figure 1 and 2), serum AEA levels were significantly increased in pale ear mice as early as 8 days after initial bleomycin exposure and remained elevated until Day 42 (Figure 5E). Furthermore, it was previously reported that expression of AEA and CB_1_R was similarly increased in the lungs of HpsPF mice 8 days after the bleomycin exposure (24). These findings suggest that activation of CB_1_R in lungs may be involved in the fibrosis initiation and progression stages in HPSPF. Unlike the pale ear mice, the bleomycin challenge did not increase circulating AEA level in wild-type mice (data not shown). This further demonstrates that circulating AEA could be a specific early prognostic blood biomarker for the fibroproliferative process in HPSPF.

**Figure 5.**
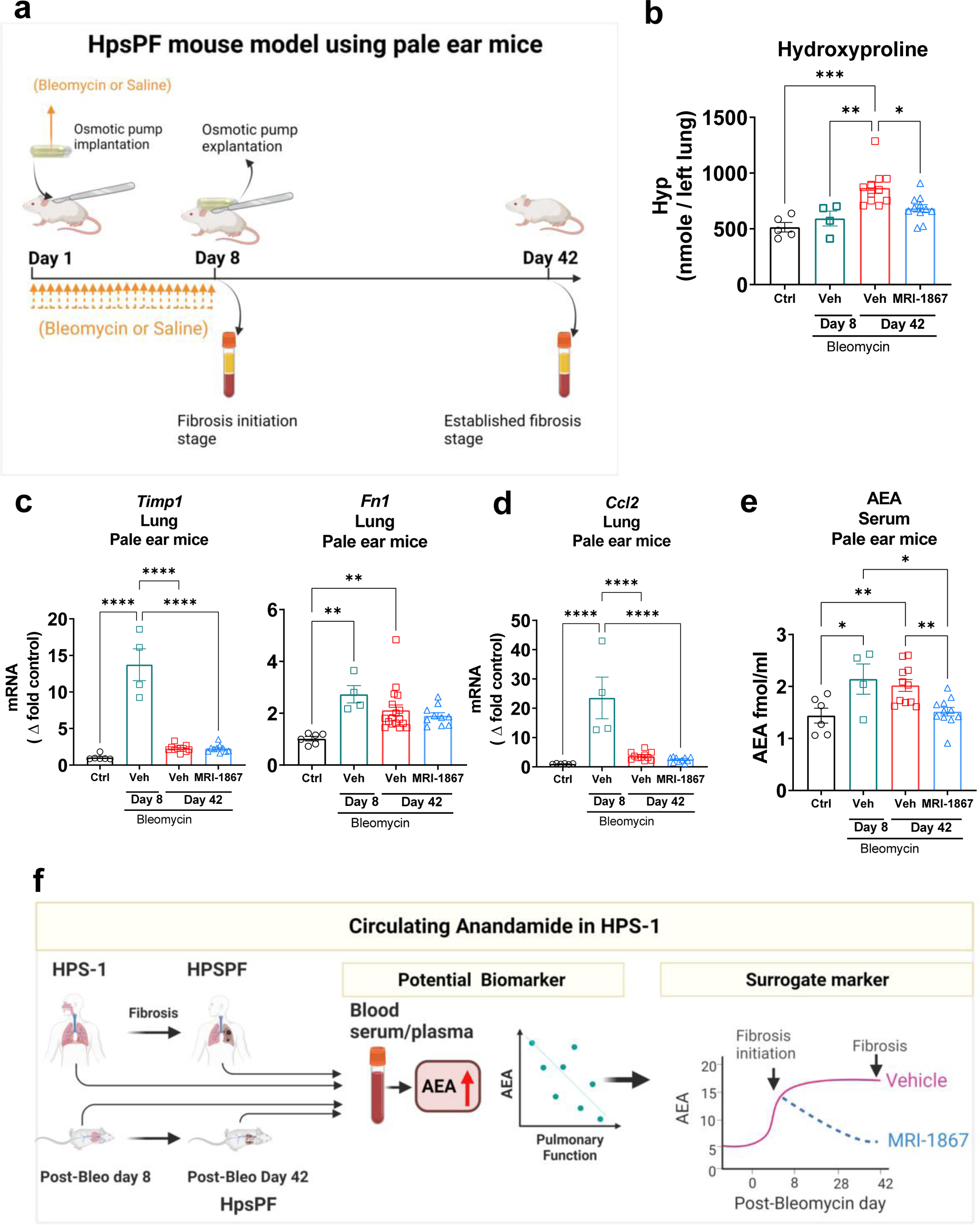
MRI-1867 treatment completely reversed bleomycin induced increase in circulating AEA level in HpsPF mouse model. (**A**) Schematic presentation of bleomycin-induced pulmonary fibrosis model in pale ear mice (HpsPF). Daily oral MRI-1867 treatment at 10 mg/kg dose started at Day 8 and continued until Day 42 post bleomycin. (**B**) Hydroxyproline levels in lungs. Gene expression of (**C**) fibrotic markers *Timp1* and *Fn1*, and (**D**) inflammatory marker *Ccl2* in lungs. (**E**) Levels of AEA in serum samples of pale ear mice. (**F**) Summary scheme depicting the circulating AEA as a biomarker and surrogate marker in HPSPF. Data represent mean + S.E.M. *n* = 6 mice for control (Ctrl, pale ear mice infused with saline instead of bleomycin), *n* = 4 HpsPF mice with bleomycin+vehicle at Day 8, *n* = 11 HpsPF mice with bleomycin+ vehicle at Day 42, *n* = 11 HpsPF mice with bleomycin+MRI-1867 at Day 42. Data were analyzed by one-way ANOVA followed by Dunnett’s multiple comparisons test. *(*P* ≤ 0.05), **(*P* ≤ 0.01).

### Increased Circulating AEA in the HpsPF Mouse Model was Completely Reversed by Peripheral CB_1_R Antagonist Zevaquenabant Treatment

CB_1_R antagonism has been identified as a therapeutic strategy for HPSPF (24), IPF (23), and radiation-induced PF (36). Recently, zevaquenabant, a clinical stage hybrid inhibitor of CB_1_R and iNOS, emerged as a candidate drug for HPSPF, since zevaquenabant treatment attenuated fibrosis progression in bleomycin-induced PF in pale ear mice (HpsPF) (24). It is important to mention that starting zevaqeunabant administration at the subclinical fibrotic stage (Day 8 post bleomycin) completely reversed the elevated circulating AEA levels (Fig. 5E) and fibrosis (Fig. 5B) in pale ear mice. As previously reported, this zevaquenabant treatment regimen also reversed the increased levels of AEA and CB_1_R expression in lungs (24). All these findings demonstrate the target engagement of zevaqeunabant for the inhibition of the endocannabinoid/CB_1_R system. Accordingly, circulating AEA levels could also be investigated as a surrogate peripheral blood marker to monitor the therapeutic efficacy of peripheral CB_1_R antagonists such as zevaquenabant in a prospective clinical trial in HPSPF (Figure 5F).

## Discussion

PF development in HPS-1 patients is universal and closely associated with age. Typically, upon onset, PF in HPS-1 patients advances rapidly, with end stage disease in HPSPF patients occurring approximately 3 years after clinical diagnosis (37). Blood biomarkers capable of accurately detecting subclinical PF and predicting onset of clinical disease are crucial for identifying patients at risk of the disease progression. Such biomarkers could be instrumental in facilitating early therapeutic interventions, potentially delaying or even preventing PF onset in these patients. Our current study indicates that circulating AEA could serve as an early disease biomarker of PF in HPS patients. Previous studies have shown that increased AEA in tissues leads to a positive feed-forward loop via overactivation of CB_1_R (38). In various cases where AEA and CB_1_R are overactivated, both genetic deletion and pharmacological inhibition of CB_1_R not only attenuated CB_1_R overactivation but also reversed pathologically increased AEA levels and provided therapeutic benefit (23, 24). In this study, treatment with the hybrid CB_1_R/iNOS antagonist zevaquenabant successfully attenuated the elevated AEA levels in the pale ear mice, confirming CB_1_R target engagement and efficacy in bleomycin-induced PF in (Figure 5). Therefore, circulating AEA could also serve as a potential surrogate marker in the clinical trials investigating peripheral CB_1_R antagonists such as zevaquenabant or monlunabant.

While HPSPF and other progressive pulmonary fibrotic diseases such as IPF share similar disease manifestations, there are notable differences between these disorders. HPS-1 PF generally affects middle-aged adults, whereas IPF impacts older adults (35). Although, pirfenidone is an approved anti-fibrotic medication for IPF, its efficacy in HPSPF remains inconclusive (29, 39, 40). Recognizing unique features in the pathological processes and pathways of these conditions, therefore, is key to optimizing anti-fibrotic efficacy and develop patient stratification strategies for therapeutic interventions for HPSPF. Notably, increased AEA levels were observed in the BALF of human IPF (25) and HPSPF (24) subjects, indicating dysregulated endocannabinoid/CB_1_R signaling involving fibrotic process in the lung microenvironment in PF. Our study found that elevated circulating AEA is a distinctive characteristic of HPS-1 patients, absent in IPF (Figure 1, 2). Therefore, the distinct observation between HPSPF and IPF in circulating peripheral AEA levels suggests that a source of AEA in serum may not be a spill over from fibrotic lung microenvironment as it is only seen in HPSPF. Another distinctive feature between HPSPF and IPF is an age dependent increase in the circulating AEA level, which was seen only in HPSPF but not in IPF patients. However, given the limited sample size of IPF patients in this study, further research is needed to evaluate endocannabinoids as blood biomarkers in IPF and other progressive fibrotic lung diseases.

Finally, our findings (Figure 1, 2) show that circulating AEA levels begin increasing with fibrotic processes in lungs even at the subclinical stages of HPSPF. Interestingly, in longitudinal HPSPF progression monitoring study, serum AEA levels almost doubled with the progression of fibrosis as evident from FVC values dropped from ∼90% predicted to ∼50% predicted values (Figure 4D-F). This trend was also observed in our preclinical study in the mouse model of HpsPF, where circulating AEA levels were significantly elevated in pale ear mice before fibrosis was evident at Day 8 post bleomycin, and remained elevated until Day 42 (Figure 5), at which point there is established fibrosis and a significant decline in PFTs (24). Since these elevated AEA levels are negatively correlated with PFTs in HPS-1 subjects (Figure 1, 2), both human and animal data support the use of circulating AEA levels as potential biomarkers for both subclinical and overt HPSPF. Taken together, these data indicate that AEA could serve as a diagnostic and prognostic biomarker for pulmonary fibrosis, including HPSPF, and warrant further investigations into endocannabinoid levels as exploratory endpoints in clinical trials targeting progressive fibrotic lung diseases.

## Data Availability

All data produced in the present study are available upon reasonable request to the authors

## Acknowledgments

This research was supported by the Intramural Research Programs of the National Institute on Alcohol Abuse and Alcoholism (NIAAA) (Z1A AA000355), the National Human Genome Research Institute (NHGRI) (Z01 HG000215-21), National Institutes of Health (NIH), and the American Thoracic Society Foundation Research Program and the Hermansky-Pudlak syndrome Network (RC). The summary and experimental schemes were generated in Biorender.com. We thank Ms. Judith Harvey-White (NIH, National Institutes of Alcohol Abuse and Alcoholism) for her technical assistance with liquid chromatography–tandem mass spectrometry experiments, and Yolanda L. Jones, NIH Library Editing Service, for editing assistance. We especially thank our patients with HPS and their families for partnering with investigators and clinicians and making this research possible.

## Author contributions

Conceptualization, R.C., W.A.G., B.R.G.; data curation, R.C., M.A, B.L.G.Z., M.X.G.Z., K.J.O., M.B. and B.R.G..; formal analysis, R.C., M.A, investigation, R.C., M.A., A.B., J.K.P, C.N.Z., K.J.O., W.J.I., M.C.V.M., and B.R.G.; methodology, R.C., M.R.I., K.J.O.B., and B.R.G.; project administration, R.C. software, M.A., and R.C.; supervision, R.C.; visualization, M.A., and R.C.; writing – original draft, R.C.; review & editing, all authors.; funding acquisition, R.C., W.A.G.

